# Phenotypic Associations and Shared Genetic Etiology between Bipolar Disorder and Cardiometabolic Traits

**DOI:** 10.1101/2020.03.31.20048884

**Authors:** Anna E. Fürtjes, Jonathan R.I. Coleman, Jess Tyrrell, Cathryn M. Lewis, Saskia P. Hagenaars

**Affiliations:** Social, Genetic and Developmental Psychiatry Centre, Institute of Psychiatry, Psychology & Neuroscience, King’s College London, London, UK; National Institute for Health Research Maudsley Biomedical Research Centre, South London and Maudsley NHS Trust, London, UK; Genetics of Complex Traits, The College of Medicine and Health, University of Exeter, The RILD Building, RD&E Hospital, Exeter, EX2 5DW

## Abstract

**Background:** People with bipolar disorder (BPD) are more likely to die prematurely, which is partly attributed to comorbid cardiometabolic traits. BPD has a bidirectional relationship with cardiometabolic traits, but their shared etiology is poorly understood. This study aimed to examine the phenotypic associations and shared genetic etiology between BPD and various cardiometabolic traits.

**Methods:** In a subset of the UK Biobank sample (*N* = 67,317) we investigated phenotypic associations between BPD (*n* = 4,155) and cardiometabolic traits, represented by biomarkers, anthropometric traits and cardiometabolic diseases. To determine the shared genetic etiology, polygenic risk scores and genetic correlations were calculated between BPD and these traits.

**Results:** Several cardiometabolic traits were significantly associated with increased risk for BPD, namely low total cholesterol, low high-density lipoprotein cholesterol, high triglycerides, high glycated haemoglobin (HbA _1c_), low systolic blood pressure, high body mass index, high waist-to-hip ratio, as well as stroke, coronary artery disease and type 2 diabetes diagnosis. Polygenic risk score regressions showed that higher polygenic risk for triglycerides, waist-to-hip ratio, coronary artery disease, and type 2 diabetes was associated with increased risk for bipolar disorder. These associations were not replicated when using genetic correlations.

**Conclusions:** This large study identified phenotypic cardiometabolic risk factors for BPD. Moreover, it found that comorbidities between BPD and waist-to-hip ratio, triglycerides, type 2 diabetes and coronary artery disease may be based on shared genetic etiology. These results provide a better understanding of the phenotypic and genetic comorbidity between BPD and cardiometabolic traits.

## Introduction

Bipolar disorder (BPD) is considered among the twenty most debilitating neuropsychiatric diseases (World Health Organization, 2008), partly because it is often accompanied by excess morbidity and premature mortality. This co-occurrence was attributed by previous studies to comorbid cardiovascular and metabolic diseases, including coronary artery disease, stroke, and type 2 diabetes, as well as risk factors such as high lipid levels in the blood and obesogenic measures (Penninx & Lange, 2018 for an overview).

There is epidemiological evidence supporting a bidirectional relationship between cardiometabolic traits and BPD. This means that cardiometabolic traits increase the risk for BPD and vice versa. Large meta-analyses established a phenotypic link between BPD and obesity, high blood pressure and abnormal lipid levels (relative risk of metabolic abnormalities = 1.58, 95% CI, 1.24-2.03) (Vancampfort, D. et al., 2015), as well as type 2 diabetes (relative risk = 1.98, 95% CI, 1.6-2.4) (Vancampfort, D., Mitchell, et al., 2016). The relationship with coronary artery disease was less clear as a meta-analysis found associations in longitudinal assessment (adjusted hazard ratio = 1.57, 95% CI, 1.28-1.93), but not in cross-sectional data (Correll et al., 2017). Nevertheless, those studies did not unravel whether cardiometabolic abnormalities are inherent properties of BPD itself, or if the comorbidities rely on environmental factors – such as medication intake or lifestyle – or genetic influences.

BPD and the cardiometabolic traits listed above have considerable hereditary components. Based on twin studies, the heritability of BPD was estimated to be 62% (Wray & Gottesman, 2012), metabolic traits were estimated around 60%, and cardiovascular traits around 40% (Polderman et al., 2015). Supporting the genetic architecture of BPD and cardiometabolic traits as highly polygenic, SNP-based heritability for BPD was estimated to be 11-25% (Stahl et al., 2019) and estimates for cardiometabolic diseases ranged from 38% for stroke (Bevan et al., 2012), 40% for coronary artery disease (Nikpay et al., 2015), and 63% for type 2 diabetes (DIAbetes Genetics Replication Meta-analysis (DIAGRAM) Consortium et al., 2014). Recent genome-wide association studies (GWAS) identified 30 distinct loci associated with BPD (Stahl et al, 2019) and many loci associated with cardiometabolic traits (eg. 304 loci for coronary artery disease (Nelson et al., 2017), 32 for stroke (Malik et al., 2018), 343 for diabetes type 2 (Mahajan et al., 2018)).

Despite evidence for the polygenicity of BPD and cardiometabolic traits, only a few studies in the existing literature – using population-based samples – investigated shared genetic etiology between them. They revealed either no associations or suggested negative associations based on approaches analysing summary statistics. For example, using linkage disequilibrium score regression (LDSC) Stahl et al. (2019) found no genetic correlations based on the most recent BPD GWAS and GWAS for cardiometabolic traits, namely body mass index, waist-to-hip ratio, type 2 diabetes, coronary artery disease, or lipid traits such as cholesterol and triglycerides. However, since this study has been published, more recent GWASs with larger samples were conducted for most of these traits. To date, the only study that systematically examined shared genetic etiology between BPD and a variety of cardiometabolic traits found reduced risk of BPD associated with *higher* polygenic risk scores. These polygenic scores were created based on summary statistics for body mass index, total cholesterol, triglycerides, waist-to-hip ratio and several other traits. In the same study LDSC did not reveal significant genetic correlations between the traits (So, Chau, Ao, Mo, & Sham, 2019).

In summary, the evidence for shared genetic etiology between BPD and cardiometabolic traits in population-based samples is scarce. Their relationships, as well as their influencing factors, are poorly understood, and to our knowledge no comprehensive phenotypic and genetic investigation of the relationship has yet been conducted using individual-level data.

This study aims to investigate the complex relationship between BPD and cardiometabolic traits on a phenotypic and genetic level in the large UK Biobank (UKB) sample. First, we hypothesise that there is an increased risk of cardiometabolic traits in BPD. We assess phenotypic associations between BPD and three categories of cardiometabolic traits, namely anthropometric risk factors, biomarkers, and cardiometabolic diseases. Second, we hypothesise that these highly polygenic traits share a genetic basis with BPD. We use two separate approaches, PRS analyses and genetic correlations, to investigate shared genetic etiology between BPD and the cardiometabolic traits considered in the phenotypic associations.

## Methods

### Sample

The UK Biobank (UKB) is a prospective cohort study including more than half a million people from across the United Kingdom (https://www.ukbiobank.ac.uk/)(Sudlow et al., 2015). It holds extensive longitudinal health-related information from physical measurements, biological samples, and questionnaires, which were collected during visits to assessment centres and online follow-ups. Two questionnaires in the UKB assessed depressive and manic symptoms. The touchscreen questionnaire assessed a “lifetime history of mood disorders” (Smith et al., 2013) which was conducted by 172,751 participants at baseline, and the second is the mental health online follow-up questionnaire (MHQ), conducted by 157,366 participants (Davis et al., 2020). In addition, hospital in-patient data, data from the national death register, and genome-wide genotyping data was available for all participants. The UKB study received ethical approval by the Research Ethics Committee. All participants signed informed consent for their data to be analysed.

### Phenotypic measures

#### Bipolar disorder (BPD)

BPD status was determined based on the combination of data from self-report questionnaires, nurse interview, and hospital episode statistics. To classify as a case, individuals had to match diagnostic codes for bipolar disorder based on ICD-10 (Supplementary Table 4), have a self-report diagnosis of mania, bipolar disorder or manic depression at nurse interview (item 20002), qualify as a case based on the baseline questionnaire about mood disorders (Smith et al., 2013), or fulfil lifetime depression and mania criteria based on the Mental Health Questionnaire (Davis, Cullen, et al., 2019).

Healthy controls did not have any mood diagnoses or self-reports and did not take antidepressant medication identified by self-reported medication use (controls adopted from Glanville et al. (2018)).

#### Blood pressure

Systolic and diastolic blood pressure was measured at baseline for every participant and corrected for antihypertensive medication based on the recommendations by Tobin, Sheehan, Scurrah, and Burton (2005). If medication intake was reported, we added 15 mmHg to systolic blood pressure and 10 mmHg to diastolic blood pressure readings. Hypertension was determined based on high systolic and diastolic blood pressure measures (systolic ≥ 140 mmHg and diastolic ≥ 90 mmHg).

#### Body mass index

Two body mass indices were available, one calculated as weight/ height (kg/ m^2^) and the other measured using electrical impedance to quantify mass. Where both measures were available, we calculated the average of the measures and excluded participants who differed between the two measures by over 4.56 standard deviations (n = 1,164) (Yaghootkar et al., 2016). If only one measure was available (*n* = 2,923), this value was used.

#### Waist-to-hip ratio

Waist-to-hip ratio was calculated as the ratio between waist and hip circumference (cm/cm). We adjusted this ratio for body mass index to represent body mass distribution in the abdominal area independent of overall body mass.

#### Biomarkers

We analysed five biomarkers from blood biochemistry: lipid levels (total cholesterol, low-density lipoprotein cholesterol (LDL cholesterol), high-density lipoprotein cholesterol (HDL cholesterol) and triglycerides) and glycated haemoglobin (HbA _1c_), as they are relevant to cardio-metabolic traits. Quality control was performed by UKB using standardised laboratory procedures (Sinnott-Armstrong et al., 2019) (http://biobank.ndph.ox.ac.uk/showcase/showcase/docs/biomarker_issues.pdf). We corrected the levels of LDL cholesterol and triglycerides for individuals taking cholesterol-lowering medication. LDL cholesterol levels were divided by 0.7 for individuals taking cholesterol medication, while triglycerides levels were divided by 0.8 (Khera et al., 2016). Supplementary Table 3 shows the cholesterol-lowering medication used.

#### Type 2 diabetes

Type 2 diabetes cases were identified based on a combination of hospital episode statistics, using both ICD-9 and ICD-10, the national death register, and self-reported questionnaire data. In order to classify as a case for type 2 diabetes, self-reported type 2 or generic diabetes status was established in the nurse interview and the touchscreen questionnaire. However, participants were only classified as cases when they reported in the questionnaire that (1) they had not been treated with insulin in the first year after diagnosis, (2) and had been diagnosed after the age of 35 years (Tyrrell, Yaghootkar, Freathy, Hattersley, & Frayling, 2013). Type 2 diabetes controls did not fulfil these criteria and did not have any other types of diabetes (individual diagnoses for inclusion and exclusion in Supplementary Tables 5 & 6).

#### Coronary artery disease

Participants who were registered in the hospital in-patient data or the death register to have had ischemic heart diseases, or participants who had coronary revascularisation operations were classified as coronary artery disease cases in this study (individual ICD-9 & 10 diagnoses and operation codes in Supplementary Tables 7 & 8). If participants self-reported those conditions in the nurse interview or the touchscreen questionnaire, they were also considered to have coronary artery disease. Coronary artery disease controls did not fulfil those criteria.

#### Stroke

Stroke status was determined using stroke diagnoses in the hospital in-patient data and the death register. In addition, participants who self-reported to have had stroke in the nurse interview or in the touchscreen questionnaire were included as stroke cases (Schnier, Bush, Nolan, & Sudlow, 2017). Stroke controls had no diagnosed or self-reported stroke.

All items included for the binary traits (type 2 diabetes, coronary artery disease and stroke) were considered at maximum three available time points in the UKB dataset. For a summary of items and codes used for the definitions described above, refer to Supplementary Table 1.

**Table 1:**
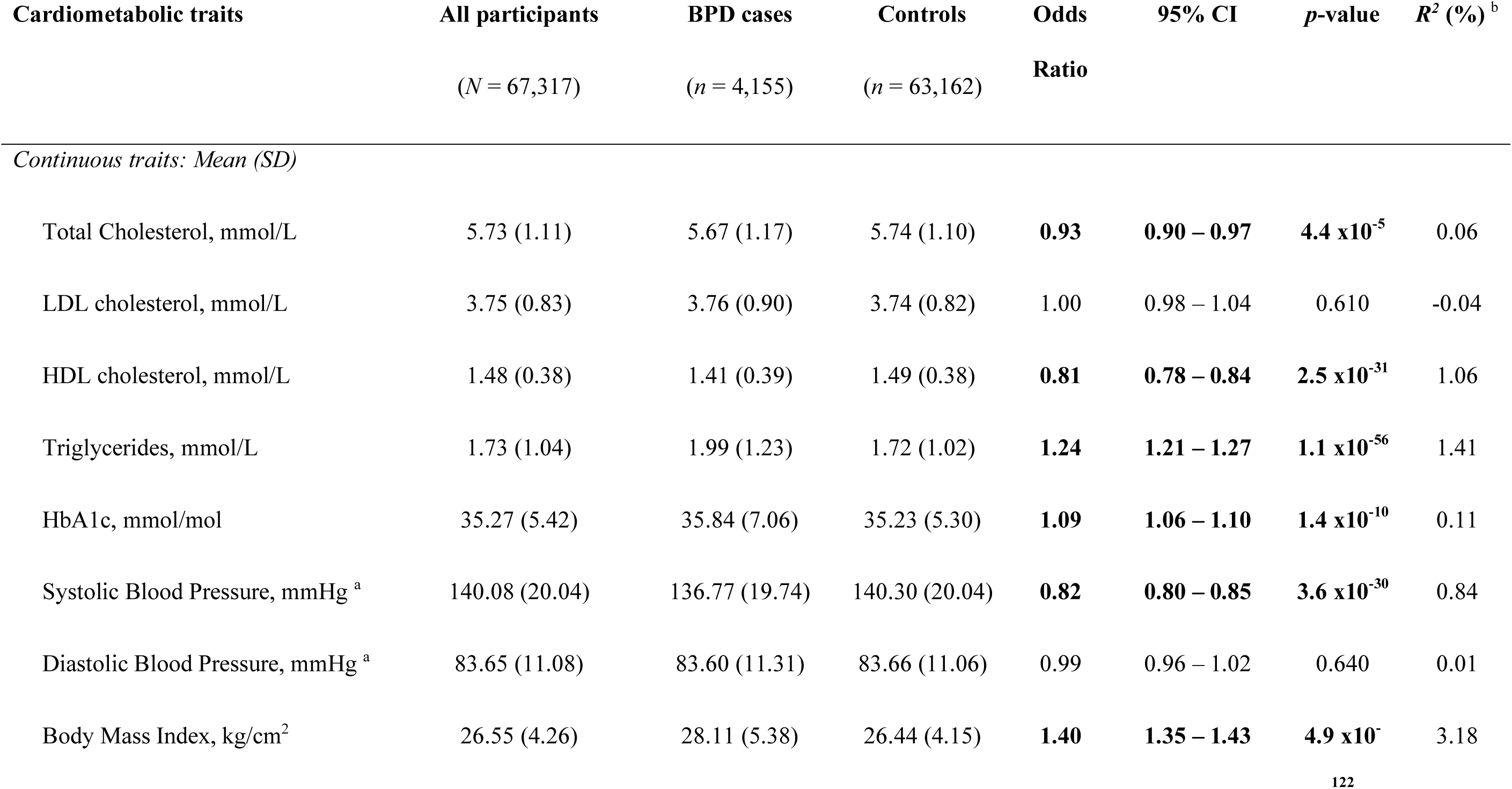
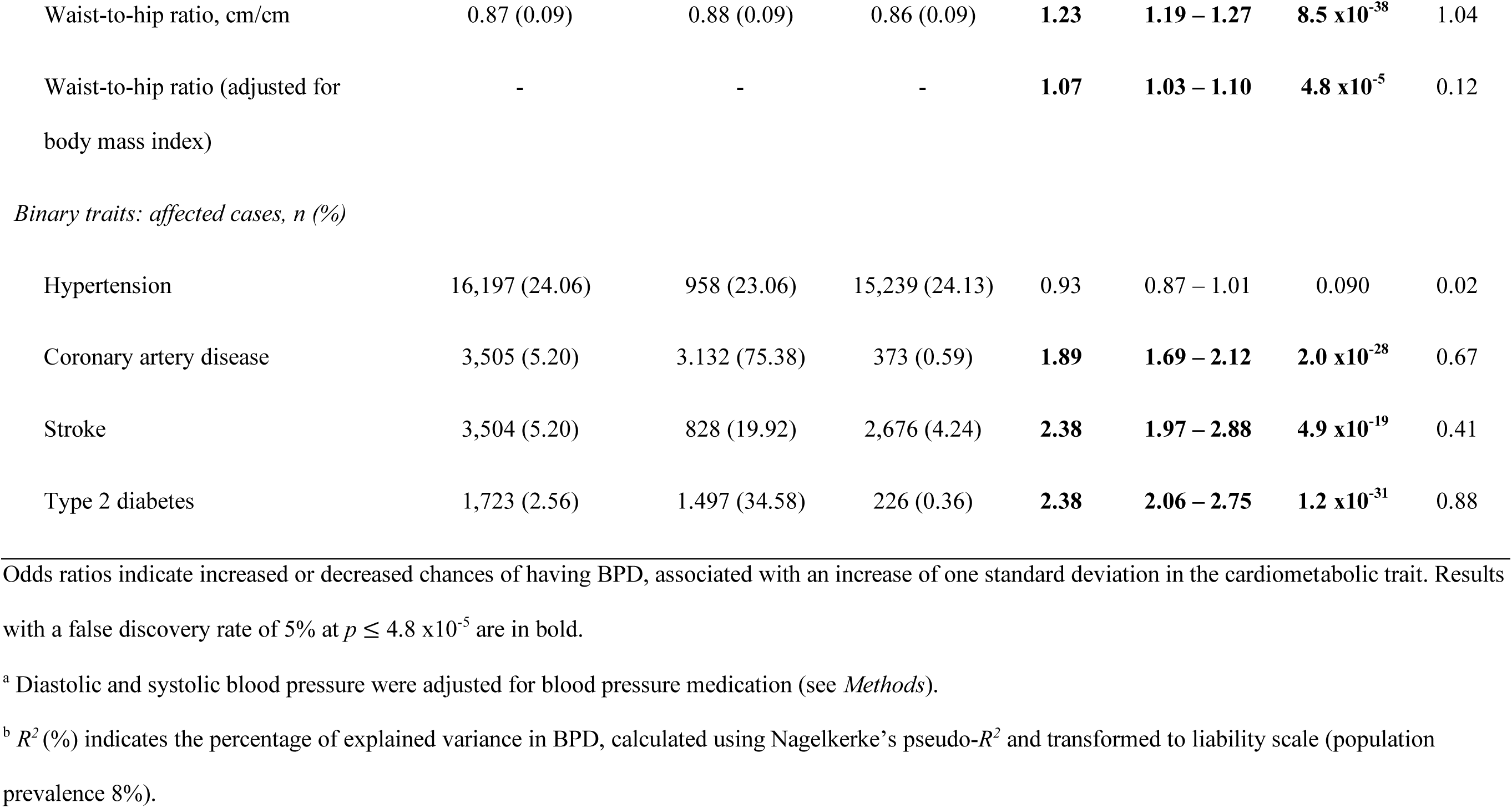
Descriptive statistics of the sample and phenotypic associations between BPD and cardiometabolic traits.

### Genotype quality control

Prior to release, UKB genetic data was genotyped on the UK BiLEVE and the UK Biobank Axiom array and underwent centralised quality control resulting in samples with 805,426 genotype markers for 488,377 available participants (Bycroft et al., 2017). Genotype data used in this study excluded rare variants (minor allele frequency < 0.01). Further quality control has been described elsewhere (Coleman et al., 2019; Hagenaars et al., 2019). Briefly, participants were excluded from analyses if they had withdrawn consent, or if they had uncommon levels of heterozygosity or high missingness (> 0.05). Participants with low call rates for genotyped SNPs (< 98%) were removed, as well as those who were related to another participant in the dataset (KING statistic of r < 0.044) (Manichaikul et al., 2010). To minimise exclusion, relatives were removed based on a greedy algorithm (eg. by excluding the child in a mother-father-child trio). Participants with mismatches between phenotypically and genetically indicated sex were removed (X-chromosome homozygosity (F _X_) > 0.5 for phenotypic females, F _X_ < 0.9 for phenotypic males). Only participants of European ancestries were analysed, which was determined based on 4-means clustering on the first two genetic principal components (Warren et al., 2017). Within this subset, six genetic principal components were identified to adjust for population stratification and technical artefacts in downstream analyses (Abraham, Qiu, & Inouye, 2017).

### Statistical analyses

#### Phenotypic associations

In order to assess phenotypic relationships between BPD status (*n* _*cases*_ = 4,155; n _controls_ = 63.162) and cardiometabolic traits, we performed logistic regressions in R v3.5.0 (R Core Team, 2013). The following continuous traits were standardised prior to analysis: total cholesterol, LDL and HDL cholesterol, triglycerides, HbA _1c_, diastolic and systolic blood pressure, body mass index and waist-to-hip ratio, unadjusted and adjusted for body mass index. All the models were adjusted for assessment centre. In addition, the models including total cholesterol, LDL cholesterol and HDL cholesterol, triglycerides and HbA _1c_ were also adjusted for fasting time.

#### Phenotypic sensitivity analyses

(1) To test whether adjustment for blood pressure medication influences our associations, we recalculated these associations excluding individuals whose blood pressure had been adjusted for medication (*N* = 11,172). (2) In order to rule out sex effects, we performed a post-hoc re-analysis of the phenotypic associations adjusting for sex. (3) Additionally, we re-calculated the phenotypic associations with a more conservative definition of BPD, only including BPD cases based on hospital-in patient data, thereby probably reflecting more severe cases (*n* _*cases*_ *= 920; n* _*controls*_ *= 63,162*).

#### Polygenic risk score (PRS) regressions

To test for shared genetic etiology between BPD and cardiometabolic traits, we created polygenic risk scores, representing genetic propensity to cardiometabolic traits for every participant. PRSs were created for total cholesterol, LDL and HDL cholesterol, triglycerides (Willer et al., 2013), HbA _1c_ (Wheeler et al., 2017), diastolic and systolic blood pressure (Evangelou et al., 2018), body mass index (Locke et al., 2015) and waist-to-hip ratio, unadjusted and adjusted for body mass index (Shungin et al., 2015), coronary artery disease (Nikpay et al., 2015), stroke (Malik et al., 2018), and type 2 diabetes (Scott et al., 2017) (availability of summary statistics in Supplementary Table 9). The summary statistics did not include data from UK Biobank, as PRS including sample overlap could lead to biased estimates. Hypertension was not included in the PRS analyses as there are no available GWAS summary statistics. Every PRS was created at eleven *p*-value thresholds (PRS _PT **=**_ 5×10^−8^, 1×10^−5^, 0.001, 0.01, 0.05, 0.1, 0.2, 0.3, 0.4, 0.5, 1). PRSice v2 (https://github.com/choishingwan/PRSice) (Choi, Heng Mak, & O’Reilly, 2018; Euesden, Lewis, & O’Reilly, 2015) was used to perform clumping –within windows of 250 kilobases (r^2^ < 0.25) – and to create the PRSs.

Shared genetic etiology between BPD and cardiometabolic traits was investigated using logistic regression with the standardised cardiometabolic trait PRSs, performed at each p-value threshold. The regression models were adjusted for assessment centre, genotyping batch, and six genetic principal components to control for population stratification.

#### Sensitivity analyses for PRS associations

(1) We aimed to identify post-hoc whether significant PRS associations found in the primary analysis were driven by phenotypic associations between BPD and binary cardiometabolic traits. To this end, modelling was repeated excluding cases affected by the binary cardiometabolic trait under investigation (excluded type 2 diabetes cases = 1,723; excluded coronary artery disease cases = 3,505). Relative attenuation in *R*^*2*^ was calculated as the difference between original *R*^*2*^ and the sensitivity *R*^*2*^, divided by the original *R*^*2*^. (2) PRS regressions were re-calculated including sex as a covariate. (3) Moreover, PRS regressions were re-assessed employing a more conservative definition of BPD, including BPD cases only based on hospital-in patient.

Explained variances in the primary phenotypic and PRS analysis (*n* _*cases*_ = 4,155) were calculated using Nagelkerke’s pseudo-*R*^*2*^ and transformed to liability scale using a population prevalence of 8%, which was suggested for screening with BPD questionnaires (Cerimele, Chwastiak, Dodson, & Katon, 2014). A population prevalence of 1% was used for analyses considering cases with hospital-in patient diagnoses only (Pini et al., 2005).

#### Genetic correlations

Genetic correlations were calculated between BPD and cardiometabolic traits using linkage disequilibrium score regression (LDSC) with the default HapMap LD reference (https://github.com/bulik/ldsc) (Bulik-Sullivan et al., 2015). We used the most recent GWAS summary statistics for BPD (Stahl et al., 2019), total cholesterol, LDL cholesterol, HDL cholesterol, triglycerides (Willer et al., 2013), HbA _1c_ (Wheeler et al., 2017), diastolic and systolic blood pressure (Evangelou et al., 2018), body mass index (Yengo et al., 2018) and waist-to-hip ratio, unadjusted and adjusted for body mass index (Shungin et al., 2015), coronary artery disease (Nikpay et al., 2015), stroke (Malik et al., 2018), and type 2 diabetes (Scott et al., 2017) (availability of summary statistics in Supplementary Table 9). Genetic correlations calculated with LDSC are robust to sample overlap, so summary statistics including the UKB sample could be included in these analyses.

#### Correction for multiple testing

To correct for multiple testing, we used the Benjamini-Hochberg false discovery rate method (Benjamini & Hochberg, 1995) for every separate analysis described above, at a false discovery rate of 0.05. *P*-values below or equal to 4.8 x10^−5^ were considered significant in the phenotypic analyses, and *p*-values below or equal to 8.6 x10^−3^ were considered significant in the PRS analyses.

## Results

The sample in this study consisted of 67,317 participants (49% females), with a mean age of 57 years (*SD* = 7.65). The sample included 4,155 BPD cases and 63,162 unaffected controls. Descriptive statistics of cardiometabolic traits by BPD case control status are shown in Table 1 (for descriptive statistics stratified by sex refer to Supplementary Table 2).

**Table 2.**
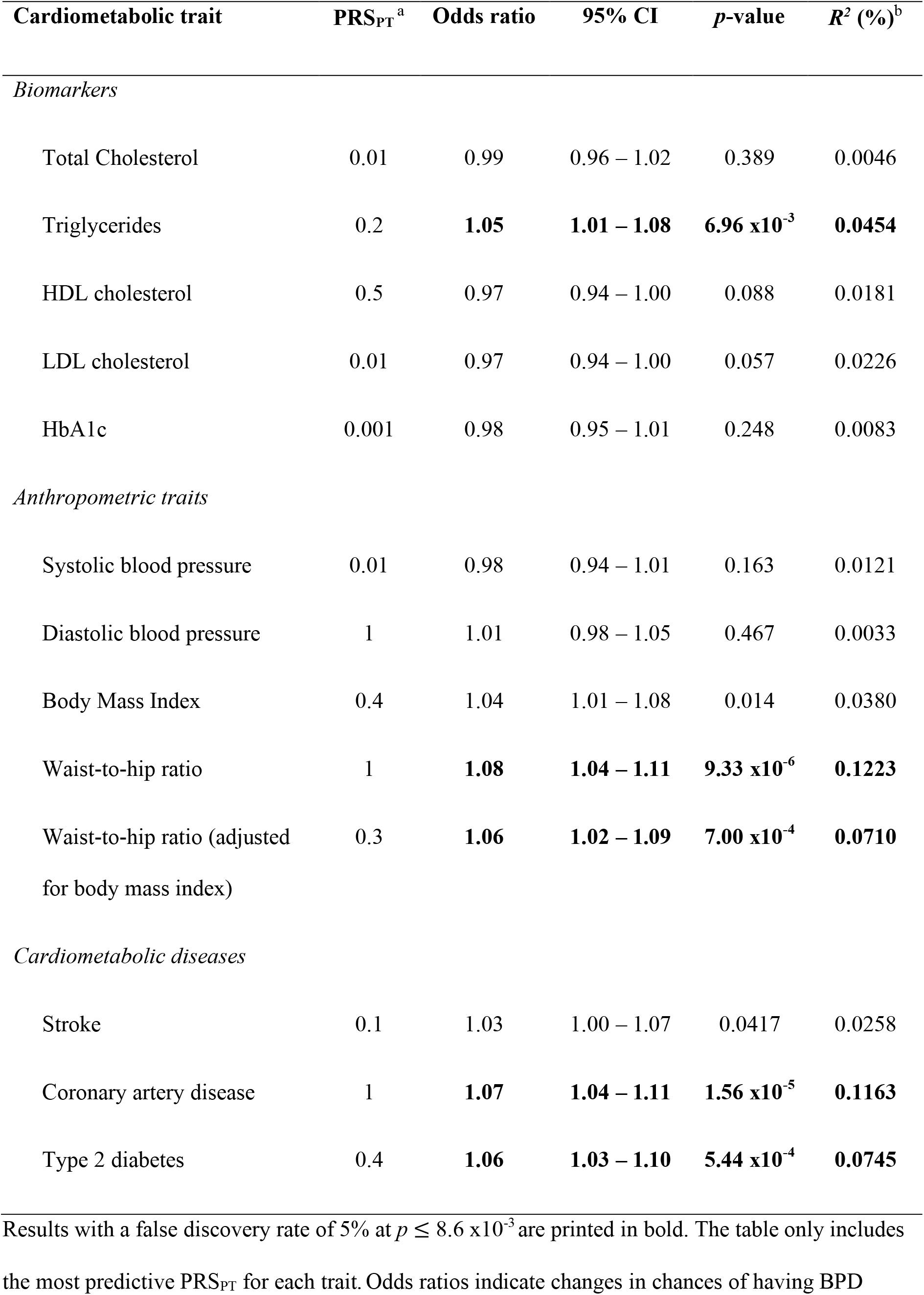
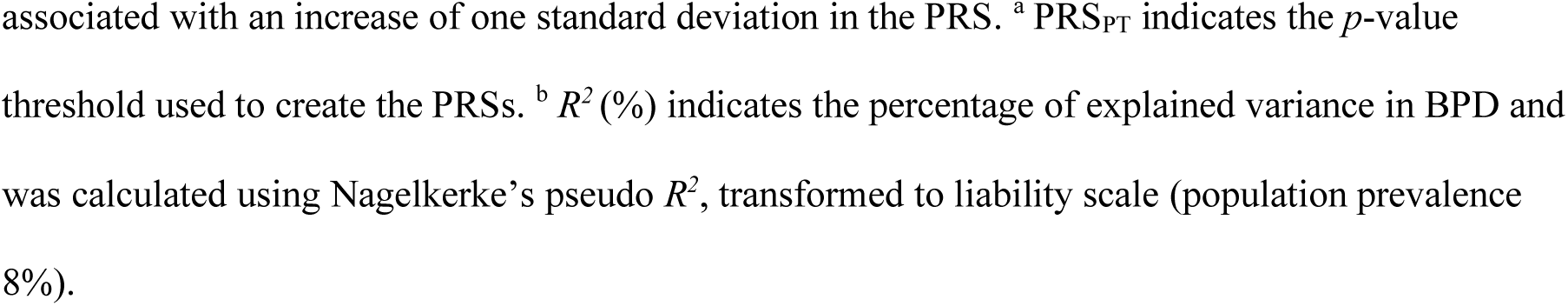
Associations between BPD and PRS for each cardiometabolic traits at the most predictive threshold.

| Cardiometabolic trait | PRS<sub>PT</sub><sup>a</sup> | Odds ratio | 95% CI | p-value | R<sup>2</sup> (%)<sup>b</sup> |
| --- | --- | --- | --- | --- | --- |
| *Biomarkers* |  |  |  |  |  |
| Total Cholesterol | 0.01 | 0.99 | 0.96 – 1.02 | 0.389 | 0.0046 |
| Triglycerides | 0.2 | **1.05** | **1.01 – 1.08** | **6.96 x10<sup>-3</sup>** | **0.0454** |
| HDL cholesterol | 0.5 | 0.97 | 0.94 – 1.00 | 0.088 | 0.0181 |
| LDL cholesterol | 0.01 | 0.97 | 0.94 – 1.00 | 0.057 | 0.0226 |
| HbA1c | 0.001 | 0.98 | 0.95 – 1.01 | 0.248 | 0.0083 |
| *Anthropometric traits* |  |  |  |  |  |
| Systolic blood pressure | 0.01 | 0.98 | 0.94 – 1.01 | 0.163 | 0.0121 |
| Diastolic blood pressure | 1 | 1.01 | 0.98 – 1.05 | 0.467 | 0.0033 |
| Body Mass Index | 0.4 | 1.04 | 1.01 – 1.08 | 0.014 | 0.0380 |
| Waist-to-hip ratio | 1 | **1.08** | **1.04 – 1.11** | **9.33 x10<sup>-6</sup>** | **0.1223** |
| Waist-to-hip ratio (adjusted for body mass index) | 0.3 | **1.06** | **1.02 – 1.09** | **7.00 x10<sup>-4</sup>** | **0.0710** |
| *Cardiometabolic diseases* |  |  |  |  |  |
| Stroke | 0.1 | 1.03 | 1.00 – 1.07 | 0.0417 | 0.0258 |
| Coronary artery disease | 1 | **1.07** | **1.04 – 1.11** | **1.56 x10<sup>-5</sup>** | **0.1163** |
| Type 2 diabetes | 0.4 | **1.06** | **1.03 – 1.10** | **5.44 x10<sup>-4</sup>** | **0.0745** |

Results with a false discovery rate of 5% at $p \leq 8.6 \times 10^{-3}$ are printed in bold. The table only includes the most predictive PRS<sub>PT</sub> for each trait. Odds ratios indicate changes in chances of having BPD
associated with an increase of one standard deviation in the PRS. <sup>a</sup> PRS<sub>PT</sub> indicates the *p*-value threshold used to create the PRSs. <sup>b</sup> $R^2$ (%) indicates the percentage of explained variance in BPD and was calculated using Nagelkerke's pseudo $R^2$ , transformed to liability scale (population prevalence 8%).

**Table 3.** Genetic correlations between BPD and 13 cardiometabolic traits calculated using LDSC regression

| Cardiometabolic trait | $h^2$ | $h^2$ se | $r_g$ | 95% CI | $p$-value |
| --- | --- | --- | --- | --- | --- |
| *Biomarkers* |  |  |  |  |  |
| Total Cholesterol | 0.2472 | 0.0384 | 0.016 | -0.044 – 0.076 | 0.611 |
| Triglycerides | 0.2857 | 0.0620 | 0.008 | -0.052 – 0.067 | 0.805 |
| HDL cholesterol | 0.2474 | 0.0388 | 0.010 | -0.039 – 0.060 | 0.681 |
| LDL cholesterol | 0.2209 | 0.0466 | 0.013 | -0.047 – 0.073 | 0.673 |
| HbA1c | 0.0627 | 0.0088 | -0.013 | -0.092 – 0.066 | 0.747 |
| *Anthropometric traits* |  |  |  |  |  |
| Body mass index | 0.2163 | 0.0070 | -0.064 | -0.108 – -0.020 | 0.004 |
| Waist-to-hip ratio | 0.1047 | 0.0067 | 0.024 | -0.035 – 0.084 | 0.423 |
| Waist-to-hip ratio (adjusted for body mass index) | 0.1207 | 0.0081 | 0.050 | 0.003 – 0.098 | 0.038 |
| Systolic blood pressure | 0.1413 | 0.0065 | 0.024 | -0.023 – 0.071 | 0.314 |
| Diastolic blood pressure | 0.1413 | 0.0065 | 0.024 | -0.023 – 0.071 | 0.314 |
| *Cardiometabolic diseases* |  |  |  |  |  |
| Stroke | 0.0121 | 0.0015 | 0.031 | 0.064 – 0.126 | 0.517 |
| Coronary artery disease | 0.0681 | 0.0053 | -0.023 | -0.088 – 0.041 | 0.475 |
| Type 2 diabetes | 0.0792 | 0.0061 | -0.054 | -0.124 – 0.017 | 0.134 |

### Phenotypic associations

The phenotypic relationship between BPD and cardiometabolic traits was assessed using logistic regression (Table 1). Meeting multiple testing correction of *p* ≤ 4.8 x10^−5^ using a false discovery rate of 5%, we observed positive associations between BPD and the continuous cardiometabolic traits triglycerides, HbA1c, body mass index, waist-to-hip ratio, waist-to-hip ratio adjusted for body mass index, stroke, type 2 diabetes, and coronary artery disease (odds ratios in Table 1, Figure 1). Total cholesterol, HDL cholesterol, and systolic blood pressure were negatively associated with BPD (Table 1, Figure 1).

**Figure 1.**
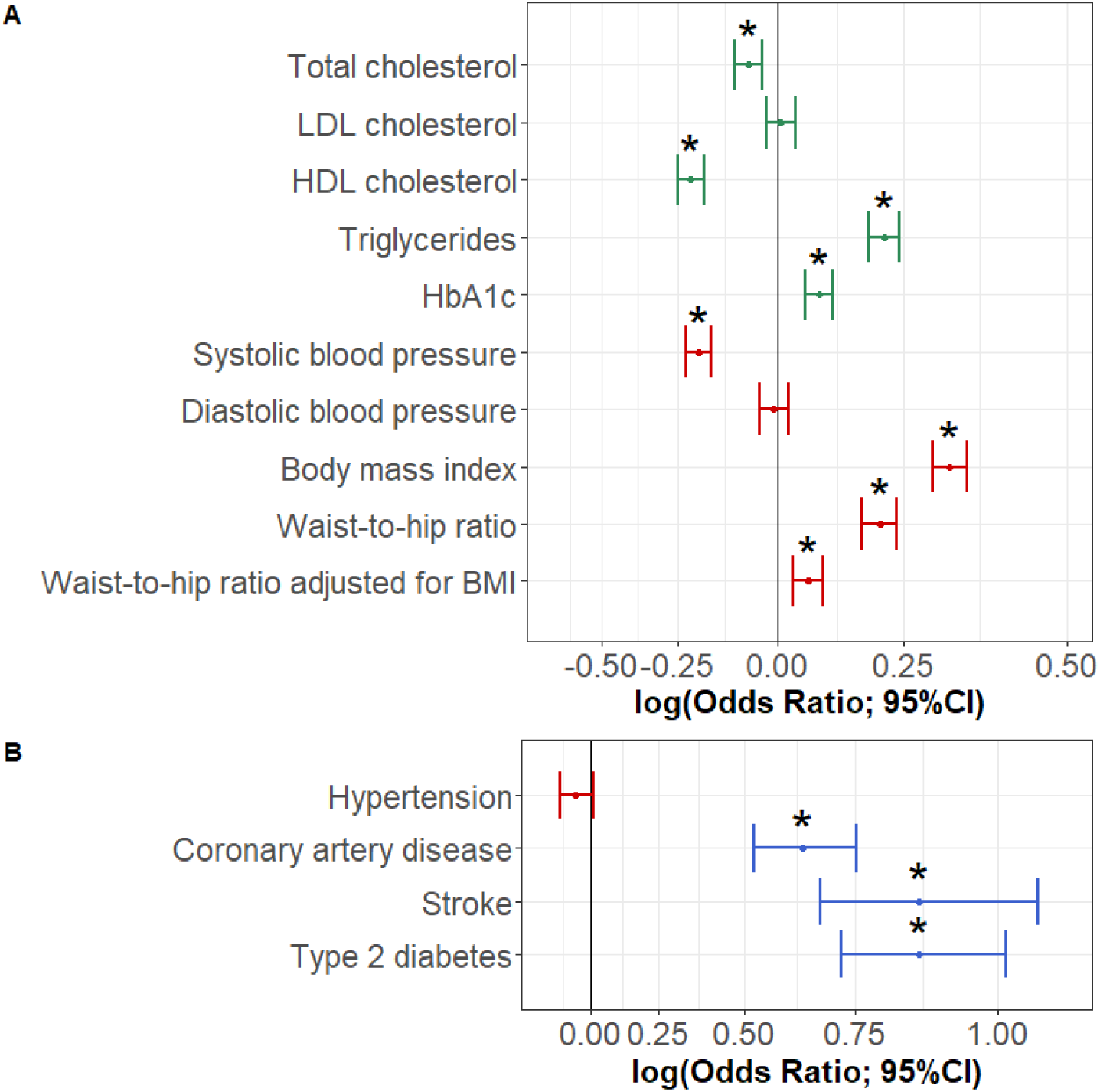
Odds ratios indicating the phenotypic associations between BPD and cardiometabolic traits. Significant associations at *p* ≤ 4.8 x10^−5^ are marked with *. Biomarkers are coloured green, anthropometric traits red and diseases blue. (A) indicates odds ratios for the continuous traits and (B) odds ratios for the binary traits.

### Phenotypic sensitivity analysis

(1) To analyse whether phenotypic associations between BPD and the three blood pressure traits were biased by the blood pressure medication adjustment, phenotypic associations were re-calculated excluding participants on blood pressure medication. Systolic blood pressure remained negatively associated (*OR* = 0.78, 95% CI, 0.75 – 0.81). Diastolic blood pressure (*OR* = 0.97, 95% CI, 0.94 – 1.01) and hypertension (*OR* = 0.89, 95% CI, 0.81 – 0.99) showed no effect on BPD, indicating that the results in the main analysis were not reliant on medication intake. (2) Adding sex as a covariate gave results consistent with those in our primary analysis (Supplementary Table 10). (3) When limiting cases to cases derived from hospital in-patient data only, odds ratios indicated larger effects (eg. stroke *OR* increased from 2.38 to 4.78), with larger amounts of variance explained (eg. coronary artery disease *R*^*2*^ increased from 0.67 to 2.41%; for all other traits refer to Supplementary Table 11).

### Polygenic risk score analysis

To test for associations between BPD and genetic liability to cardiometabolic traits, logistic regressions were performed with eleven PRSs at different *p*-value thresholds for each cardiometabolic trait. Results for the most predictive thresholds in each cardiometabolic trait are indicated in Table 2 and Figure 2. At a multiple testing correction of *p* ≤ 8.63 x10^−3^ (all traits and eleven thresholds each; FDR 5%), BPD was positively associated with PRSs for triglycerides, waist-to-hip ratio, waist-to-hip ratio adjusted for body mass index, coronary artery disease, and type 2 diabetes. The highest contribution to explaining variance in BPD was from the waist-to-hip ratio PRS (PRS _PT_ = 1) accounting for 0.13% of the variance (complete results for each PRS threshold can be found in Supplementary Tables 12).

**Figure 2.**
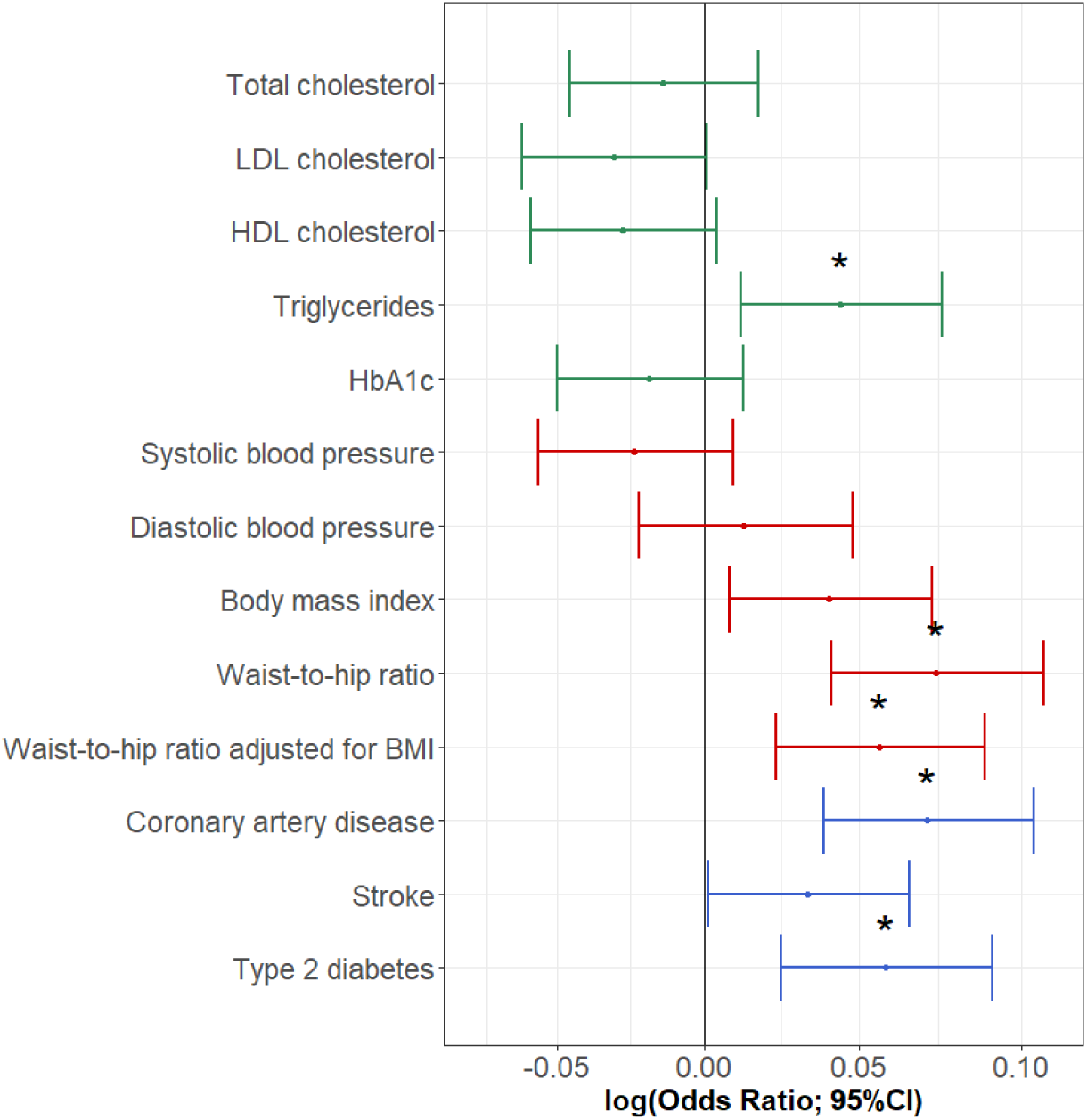
Association between BPD and the most predictive PRSs for each cardiometabolic trait. Significant associations at *p* ≤ 8.6 x10^−3^ are marked with *. Biomarkers are coloured green, anthropometric traits red and diseases blue.

### Sensitivity analyses for PRS associations

Sensitivity analyses were performed to investigate the impact by phenotypic associations between BPD and the cardiometabolic diseases on PRS associations, excluding participants affected by type 2 diabetes (1,723 cases) or coronary artery disease (3,505 cases) and re-calculating regressions for the significant PRSs. For coronary artery disease PRSs, explained variance was hardly attenuated. For example, the variance for PRS _PT_ 0.5 declined from *R*^*2*^ = 0.099% in the main analysis to *R*^*2*^ = 0.093% in the sensitivity analysis (relative attenuation across significant thresholds = −2.54 to 5.88%). For significant associations with type 2 diabetes PRSs, exclusions caused a more substantial attenuation in explained variance. For example, variance for PRS _PT_ 0.2 declined from *R*^*2*^ = 0.048 in the main analysis to *R*^*2*^ = 0.034% in the sensitivity analysis. (relative attenuation across significant thresholds 23.00 to 29.59%; Supplementary Table 12). When re-calculating the regressions adding sex as a covariate, results were consistent with the primary PRS analysis (Supplementary Table 13). To test whether the strength of the phenotype is influencing our associations, we restricted to BPD cases based on hospital in-patient diagnoses only (N _cases_ = 920). While these associations did not survive correction for multiple testing, the amount of variance explained increased across the PRSs for total cholesterol, HDL cholesterol, HbA1c and systolic blood pressure, indicating a better model fit when restricting to a more stringent phenotype (Supplementary Table 14).

### Genetic correlations

Genetic correlations between BPD and 13 cardiometabolic traits were all low and were not significant after correcting for multiple testing (Table 3, Figure 3). Body mass index (*r* _*g*_ = −0.064, *p* = 0.004) and waist-to-hip ratio adjusted for body mass index (*r* _*g*_ = 0.050, *p* = 0.038) were nominally correlated but did not survive correction for multiple testing.

**Figure 3.**
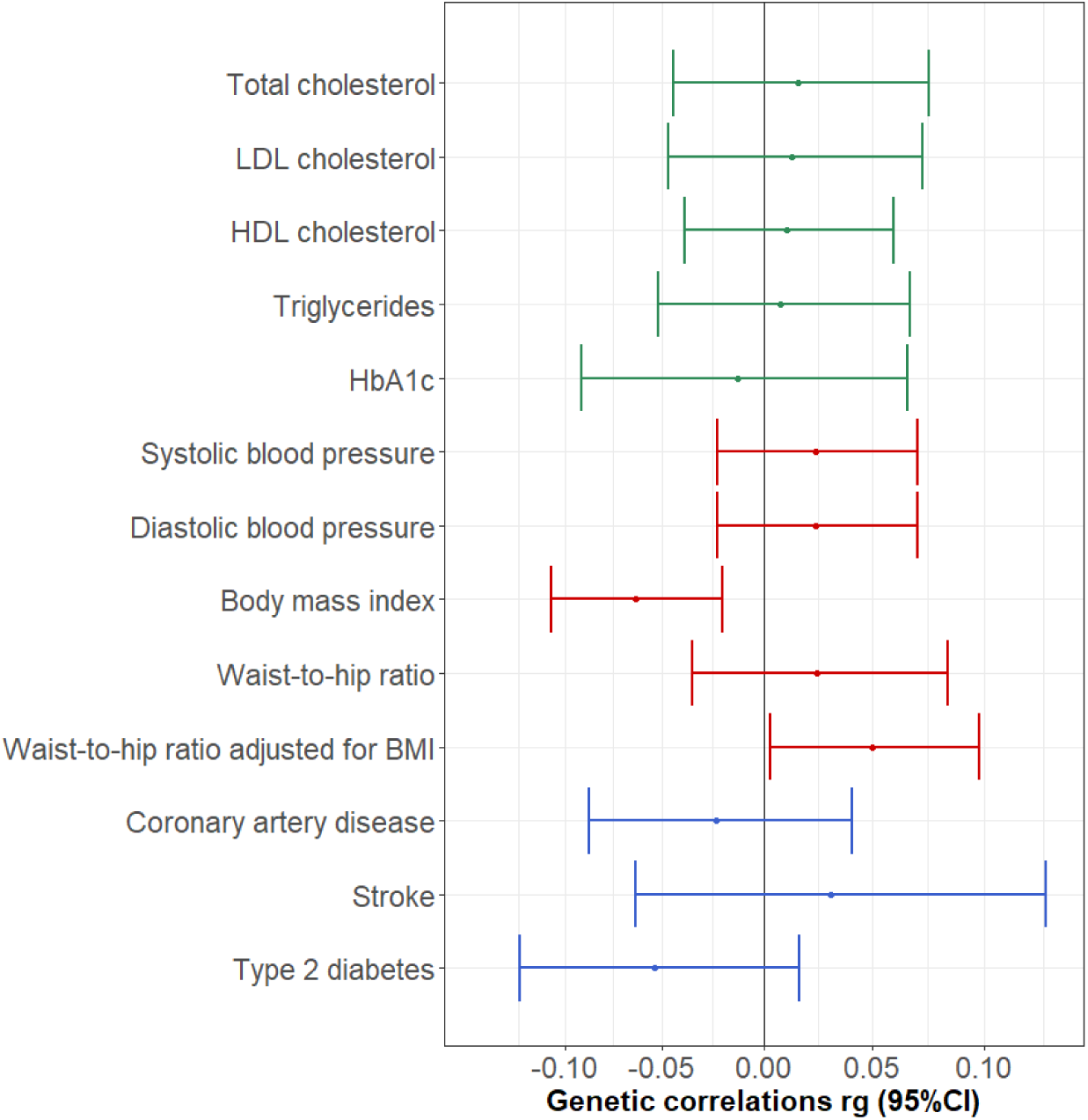
Genetic correlations between BPD and cardiometabolic traits. Biomarkers are coloured green, anthropometric traits red and diseases blue.

## Discussion

The aims of this study were to investigate the relationship between BPD and cardiometabolic traits in the UK Biobank sample on a phenotypic level and on a genetic level, using PRS analyses and genetic correlations.

The phenotypic results revealed significant associations and relatively small effect sizes with a number of cardiometabolic risk factors, namely low total cholesterol, high triglycerides, low HDL cholesterol, low systolic blood pressure, high body mass index, high waist-to-hip ratio, as well as stroke, type 2 diabetes and coronary artery disease status. These findings were largely consistent with our hypothesis that cardiometabolic risk factors and diseases are associated with an increased risk of BPD. Sensitivity analyses including a smaller subpopulation of more severe BPD cases (*N* _*cases*_ = 920) confirmed these associations and produced increased amount of variance explained, which suggests that more severe cases are more likely to demonstrate cardiometabolic abnormalities.

We discovered shared genetic etiology based on PRS associations for four cardiometabolic traits, which were triglycerides, waist-to-hip ratio, coronary artery disease and type 2 diabetes. This indicates that the phenotypic associations between these traits and BPD are at least partly based on shared genetic predisposition. Sensitivity analyses using a subpopulation of more severe BPD cases (n _cases_ = 920) failed to produce significant results, but effect sizes were consistent with effect sizes obtained in the original analysis. With the altered BPD definition, the number of cases might have been too small to capture significant PRS associations, which underlines the importance of future large-scale studies.

Our results show increased triglycerides, decreased HDL cholesterol and increased HbA _1c_ were associated with higher risk for BPD, which was consistent with meta-analyses reporting increased metabolic abnormalities in BPD (Vancampfort, D. et al., 2015; Vancampfort, D et al., 2013). It was surprising that total cholesterol in the BPD group was lowered, because we hypothesised that it acts as a risk factor for cardiometabolic disease. The existing literature provides inconsistent results, as it suggests either no effect of peripheral total cholesterol on BPD (Garcia-Portilla et al., 2009), or reduced total cholesterol in very small samples (Atmaca, Kuloglu, Tezcan, Ustundag, & Bayik, 2002). To our knowledge, this is the first large study to support evidence for a negative association.

In the PRS analysis, genetic liability to high triglyceride levels was the only biomarker that increased risk of BPD. This relationship had only been reported in an Amish pedigree with highly related individuals (Kember et al., 2018). Future studies based on larger GWAS studies for cholesterol measures might be more powerful to detect an association. However, the association with triglycerides was not replicated using genetic correlations which is also the case for the other significant PRS associations in this study. As genetic correlations are calculated with GWAS summary statistics and PRS analyses rely on individual level genotype data, we suggest that PRS analyses might have been more powerful in detecting the associations. The different results between the two methods might also reflect discontinuity between the phenotypes captured in UKB compared with PGC and findings in this study might only describe BPD relationships specific to the UKB sample instead of BPD in the general population.

We also found support for phenotypic associations between BPD and obesogenic measures, which were waist-to-hip ratio and body mass index. The latter contributed the strongest association among all traits and corroborated previous reports indicating a positive association (Correll et al., 2017). This is supported by a recent study that identified an association between excessive weight gain and brain volume loss in BPD (Bond et al., 2019). Such brain changes could imply a potential mechanism linking obesogenic traits with BPD.

In addition, genetic liability for an increased waist-to-hip ratio was associated with increased odds of having BPD, indicating shared genetic etiology. PRSs for waist-to-hip ratio unadjusted and adjusted for body mass index produced similar associations, presumably because the phenotypes used in the discovery GWAS were highly correlated (*r* = 0.82–0.95; Shungin et al., 2015). Those associations underline that waist-to-hip ratio is an important obesogenic risk factor for BPD, and that not only genetic variants associated with body fat distribution (waist-to-hip ratio) but also abdominal obesity independent of body fat (waist-to-hip ratio adjusted for body mass index) play a role in BPD pathology. Waist-to-hip ratio conveys a distinct increased risk, beyond the risk contributed by overall adiposity (indicated by body mass index for example), for cardiometabolic diseases (Canoy, 2008; Wang, Rimm, Stampfer, Willett, & Hu, 2005), mortality (Pischon et al., 2008), and BPD, as this study showed.

This study showed that diagnosis of coronary artery disease, stroke or type 2 diabetes was associated with at least two-fold increase in odds for having BPD, which is consistent with previous epidemiological findings in large meta-analyses (Correll et al., 2017; Vancampfort, D., Mitchell, et al., 2016). The PRS analyses showed that participants with a higher genetic propensity to coronary artery disease were more likely to have BPD. This relationship was consistent with our phenotypic findings, and previously reported longitudinal assessments (Correll et al., 2017). To assess whether phenotypic associations between the two traits drove the PRS association, we performed sensitivity analyses, excluding 3,505 coronary artery disease cases. The relative attenuation in explained variance between original and sensitivity analyses clustered around zero, suggesting little reduction in predictive value of the PRSs. This suggests that the shared genetic etiology with coronary artery disease was likely based on common genetic effects.

Increased genetic liability to type 2 diabetes was associated with elevated chances to have BPD. This finding was consistent with the well-established phenotypic associations between type 2 diabetes and BPD (Vancampfort, D., Correll, et al., 2016). In contrast with coronary artery disease, sensitivity analyses excluding 1,723 type 2 diabetes cases resulted in a considerable relative attenuation of explained variance. This suggests that shared genetic etiology between type 2 diabetes and BPD was partly driven by their phenotypic association.

The cardiometabolic traits identified in this study to share genetic etiology with BPD were previously reported to capture overlapping genetic effects (*r* _*g*_ = 0.22 - 0.58; (Bulik-Sullivan et al., 2015; Shungin et al., 2015; Willer et al., 2013). Additionally, they were shown to be causally linked. For example, variants associated with waist-to-hip ratio and triglycerides are causally involved with coronary artery disease and type 2 diabetes (Do et al., 2013; Emdin et al., 2017). Those causal links indicate that the traits influence each other, potentially through shared biological pathways that might also be relevant to BPD.

This study did not find consistent associations between measures of blood pressure and BPD, and these results were not influenced by our adjustment for blood pressure lowering medication. Previous meta-analyses found prevalent hypertension in BPD participants (Ayerbe et al., 2018). However, these results relied on composite hypertension measures, which makes these results difficult to compare to ours.

Our findings in this study must be interpreted considering several limitations. The *possible bipolar* status definition used in the primary analysis was chosen due to its high predictive validity (Davis, Cullen, et al., 2019). However, it may outline a subpopulation with increased prevalence of BPD symptoms, that does not correspond to a clinical sample, so caution must be taken when translating our findings. Furthermore, it is difficult to generalise the effect sizes from our findings to the general population, because participants from the UKB, and in particular participants in the mental health questionnaire, are subject to a *healthy volunteer selection bias* (Adams et al., 2018; Davis, Coleman, et al., 2019; Fry et al., 2017). Systematic differences between participants and non-participants probably have introduced spurious relationships for variables that influence participation in the UKB, and the BPD cases in our study are likely not typical BPD cases (Munafò, Tilling, Taylor, Evans, & Davey Smith, 2017). It must also be considered that phenotypic associations purely represent the manifestation of BPD alongside cardiometabolic traits, and that environmental factors, such as lithium medication, influence this relationship. Phenotypic and genetic associations explain only a fraction of variance in BPD (*R*^*2*^ phenotypes ≤ 3.4%, *R*^*2*^ PRS ≤ 0.12%), and none of the associations implicate causal underlying biological mechanisms. Lastly, as highlighted above significant associations in PRS analyses were not replicated using LDSC.

This study contributes to the existing literature by showing phenotypic associations and shared genetic etiology of BPD with several cardiometabolic traits in the UKB sample. To our knowledge, this is the largest cross-sectional study investigating this relationship considering biomarkers, anthropometric traits and cardiometabolic diseases. We performed extensive phenotyping for all included traits considering clinical, and self-report data, physical measurements and biological samples. In addition, we used the most recent available GWAS summary statistics to compute polygenic risk scores and genetic correlations.

To conclude, we showed that a variety of phenotypic cardiometabolic traits act as risk factors in BPD in the UKB sample. Body mass index was most strongly associated with BPD. Moreover, we found that comorbidities between BPD and waist-to-hip ratio, triglycerides, coronary artery disease and type 2 diabetes were partly based on shared genetic etiology. Our results underline the importance of considering cardiometabolic comorbidity in the context of BPD, and shared genetic mechanisms of comorbid physical and mental disorders.

## Data Availability

UK Biobank is a publicly available controlled-access dataset available to any bona fide researcher. This study was conducted using UK Biobank data obtained under approved application 18177.

## Acknowledgement

AEF is funded by the Social, Genetic and Developmental Psychiatry Centre, King’s College London and the National Institute of Health grant R01AG054628. SPH is funded by the Medical Research Council (MR/S0151132). JT is supported by an Academy of Medical Sciences (AMS) Springboard award, which is supported by the AMS, the Wellcome Trust, GCRF, the Government Department of Business, Energy and Industrial strategy, the British Heart Foundation and Diabetes UK (SBF004\1079).

This study presents independent research supported by the National Institute for Health Research (NIHR) Biomedical Research Centre at South London and Maudsley NHS Foundation Trust and King’s College London. The views expressed are those of the author(s) and not necessarily those of the NHS, NIHR, Department of Health or King’s College London. This study was conducted using UK Biobank data obtained under approved application 18177. We thank participants and scientists involved in making the UK Biobank resource available (http://www.ukbiobank.ac.uk/).

## Notes

### Competing Interest Statement

The authors have declared no competing interest.

